# Family history of misophonia and co-occurring neuropsychiatric conditions

**DOI:** 10.64898/2026.03.13.26347988

**Authors:** Salome Alfaro, Diana Bok, Doris Chen, Thomas V. Fernandez, Emily Olfson

## Abstract

**Objective:** To characterize the familial patterns of misophonia and other commonly co-occurring neuropsychiatric conditions.

**Methods:** We examined cross-sectional survey responses from 101 probands with misophonia and their biological parents enrolled in a genetics study.

**Results:** Probands had a mean age of 24.6 ± 11.6 years (8-64 years), were predominantly female (88%), and had high rates of co-occurring neuropsychiatric conditions, including anxiety (70%), depression (38%), ADHD (31%), and OCD (25%). Among probands, 39% had a first-degree relative with misophonia, and 48% had at least one any-degree relative with misophonia. In addition, many probands had at least one first-degree relative with anxiety (65%), depression (57%), ADHD (40%), OCD (20%), and autism (13%). Comparing rates of neuropsychiatric conditions reported by parents, mothers had significantly higher rates of misophonia (29% maternal vs. 9% paternal, *p* = 0.001) and anxiety (44% maternal vs. 26% paternal, *p* = 0.02) than fathers.

**Conclusion:** These findings provide new insight into the familial patterns of misophonia and co-occurring neuropsychiatric conditions. Future research on underlying genetic and environmental factors is needed to shed light on the observed shared predispositions for misophonia and other neuropsychiatric conditions in families.

## Introduction

Misophonia is a condition characterized by decreased tolerance for and heightened reactivity to specific sensory stimuli, most commonly sounds, but also visual and other sensory triggers, that are typically ordinary and often repetitive in nature.^1,2^ These reactions are disproportionate to the intensity of the trigger sound, cause intense emotional responses (e.g., anger, panic, rage), and are frequently accompanied by physiological (e.g., increased heart rate, sweating, muscle tension) and behavioral responses (e.g., avoidance or escape behaviors).^3-5^ These symptoms can profoundly impact an individual’s quality of life, causing significant impairment in psychosocial functioning and suicidal thoughts in up to 20% of cases.^1,6^ Although formal diagnostic criteria for misophonia have yet to be fully established, the pattern of symptoms in affected individuals is notably distinct from that of other psychiatric disorders, such as anxiety or obsessive-compulsive disorder (OCD), and audiological disorders like tinnitus or hyperacusis.^3^ Research suggests that misophonia predominantly affects females^1,5,7-11^ and typically onsets around the transition from childhood to adolescence.^5,12^

Several studies suggest that misophonia clusters in families. A few studies have assessed family history of misophonia symptoms, reporting rates ranging from 8% to 54% of affected individuals having a relative with similar symptoms or sound sensitivities.^1,3,4,11,13,14^ In the largest study, Rouw and Erfanian found that 67 out of 301 participants with misophonia (22%) had relatives with similar symptoms, and these affected family members were more often female than male.^1^ Another study of 90 children and adolescents with misophonia reported misophonia symptoms in 35% of parents, 7% of siblings, 23% of grandparents, and 23% of uncles/aunts.^14^ To our knowledge, studies have not examined family history of co-occurring neuropsychiatric conditions or directly surveyed family members. Additional family history studies are needed to further characterize how misophonia and commonly co-occurring conditions segregate within families, advancing our understanding of possible genetic and environmental risk factors.

This study aimed to investigate the family history of misophonia and related neuropsychiatric conditions in a sample of 101 individuals with a self-reported diagnosis of misophonia (probands, i.e., the primary affected individual in each family) enrolled in an ongoing genetic study of families. Using cross-sectional survey responses from probands and their biological parents, we present details on the family history of misophonia and commonly co-occurring conditions. Unlike prior studies that relied exclusively on proband-reported family history, this study directly surveyed biological parents about their own diagnostic histories, providing more reliable estimates of parental misophonia and co-occurring condition status.

## Methods

### 2.1 Procedures

Participants were recruited as part of an ongoing genetic study, approved by the Institutional Review Board (IRB) at the Yale School of Medicine, following the standards of the U.S. Federal Policy for the Protection of Human Subjects (HIC: 0301024156, most recently approved by the Yale IRB on 10/27/2025). Informed consent/assent was obtained from all participants, and human subject privacy rights were observed. Probands with misophonia were recruited through various online organizations and support groups for affected individuals between December 2023 and June 2025. Probands were eligible to participate if they answered “Yes” or “Diagnosed by a clinician” to the question about misophonia in their self-reported survey. Probands, both their biological parents, and, in some cases, affected siblings, were all enrolled as participants.

### 2.2 Participants

In this family history study, we included 101 unrelated probands with a self-reported diagnosis of misophonia; only probands with both biological parents were eligible to participate. When multiple affected siblings enrolled, only the first enrolled was included in this analysis to ensure that all probands were unrelated. When available, self-report surveys completed by parents or affected siblings were used to assess their own medical histories.

### 2.3 Instruments

Each participant (or parent/guardian on behalf of younger children) completed surveys about their demographics, co-occurring conditions, family history, and current symptoms.

To evaluate misophonia severity and impairment, the Duke Misophonia Questionnaire (DMQ) was used. Our analysis focused on the four quantitative subscales of the DMQ: Affective Responses, Physiological Response, Cognitive Response, and Impairment. The overall Symptom Severity Composite Score (Severity Score) was determined by adding the Affective, Physiological, and Cognitive Response scores. Scores greater than 41 for the overall Severity Score are indicative of higher misophonia symptoms, and scores greater than 13 on the Impairment subscale (Impairment Score) are indicative of moderate to very severe impairment.^15^ For probands who omitted a single question on the combined Severity Score (5/101), scores were calculated by assigning “0” to this question. Probands omitting two or more questions on either the Severity Score (2/101) or Impairment Score (3/101) were excluded from analyses of that subscale. Mothers and fathers who checked “Unsure” about having misophonia were considered to have misophonia only if their Severity Score was > 41 (suggestive of higher symptoms) or if their Impairment Score was > 13 (suggestive of at least moderate impairment).^15^

To assess the prevalence of co-occurring conditions, participants were asked to select one of the following responses: “Yes,” “Unsure,” “Diagnosed by a clinician,” or “No” for several co-occurring neuropsychiatric conditions. Participants were considered to have a co-occurring condition if they selected “Yes” or “Diagnosed by a clinician.” Responses marked as “Unsure,” “No,” or left blank were considered not to have the condition.

### 2.4 Family history

Participants were asked to self-report on the medical (including mental and behavioral health) histories of their mother, father, siblings, and any other family members. When available, surveys completed by parents or affected siblings were used to determine the medical histories of these relatives. In cases where these were not available and for relatives of all other degrees who were not directly surveyed, medical histories were determined based on the proband’s self-report. Probands were categorized as “simplex” or “multiplex” according to their family history status. Families were considered simplex if only the proband had misophonia, and multiplex if the proband had at least one other first-degree relative with misophonia. To compare rates of misophonia and co-occurring conditions between parents, we analyzed available medical history reports completed by mothers (n=84) and fathers (n=81) of the affected probands.

### 2.5 Statistical analyses

Analyses were conducted using R (version 4.4.0). Demographics and co-occurring conditions between simplex and multiplex probands were analyzed using two-sided Fisher’s exact tests for categorical variables (e.g., sex, race, ethnicity, diagnoses) and two-sided independent t-tests for continuous variables (i.e., age and DMQ scores). Given the exploratory nature of this study and the relatively small sample size, we did not correct for multiple comparisons; results should be interpreted accordingly, and findings require replication in independent samples.

## Results

Our analyses examined survey data from 101 probands with misophonia and their relatives. Probands had a mean age of 24 years (*SD* = 11.8; range = 8-64 years) at enrollment, and were predominantly female (88.1%), white (99.0%), and non-Hispanic/non-Latino (92%) (**Table 1**). The most common self-reported co-occurring conditions in probands with misophonia were anxiety (70%), depression (38%), attention-deficit/hyperactivity disorder (ADHD) (31%),and OCD (25%) (**Table 1**).

**Table 1.**
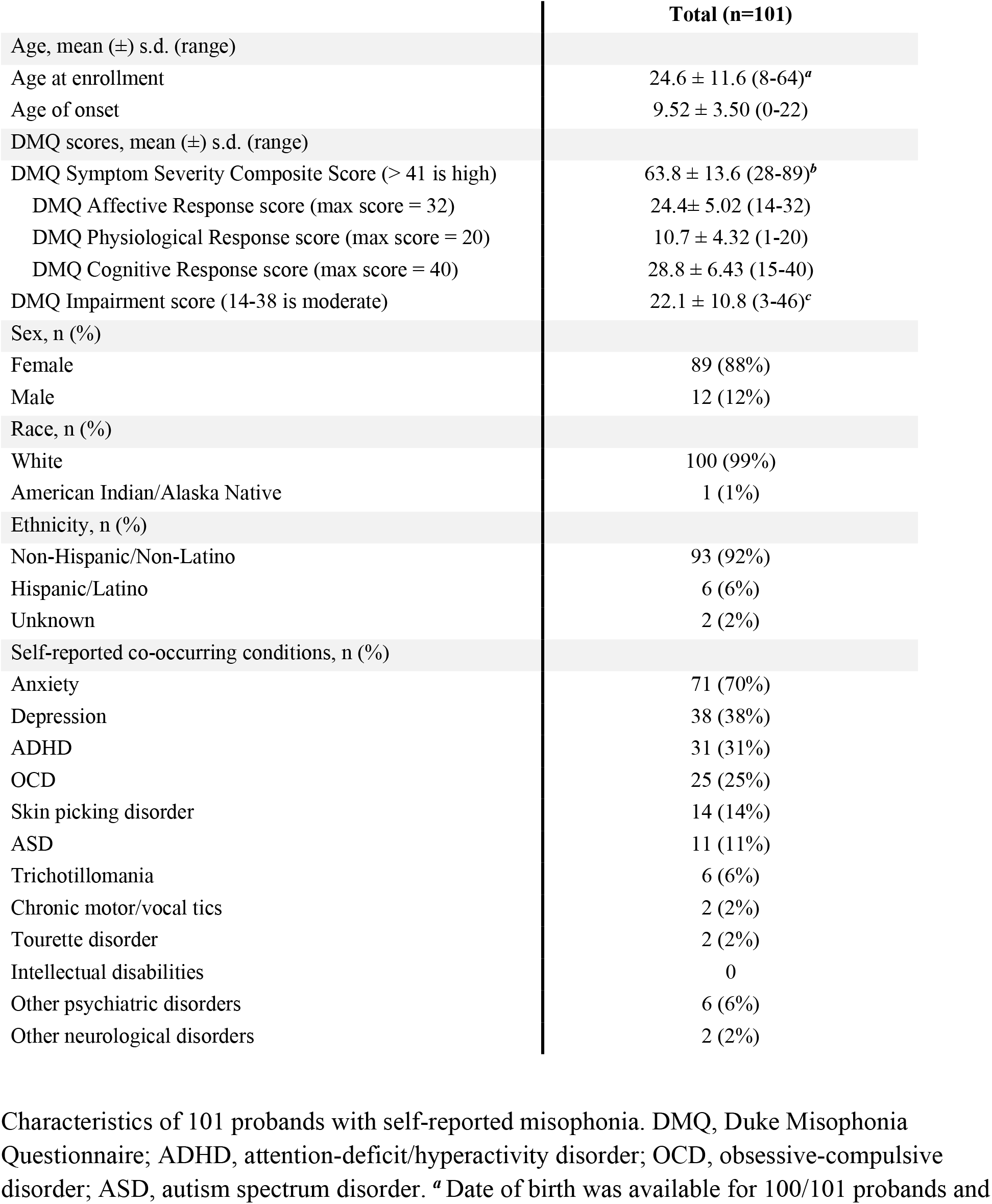
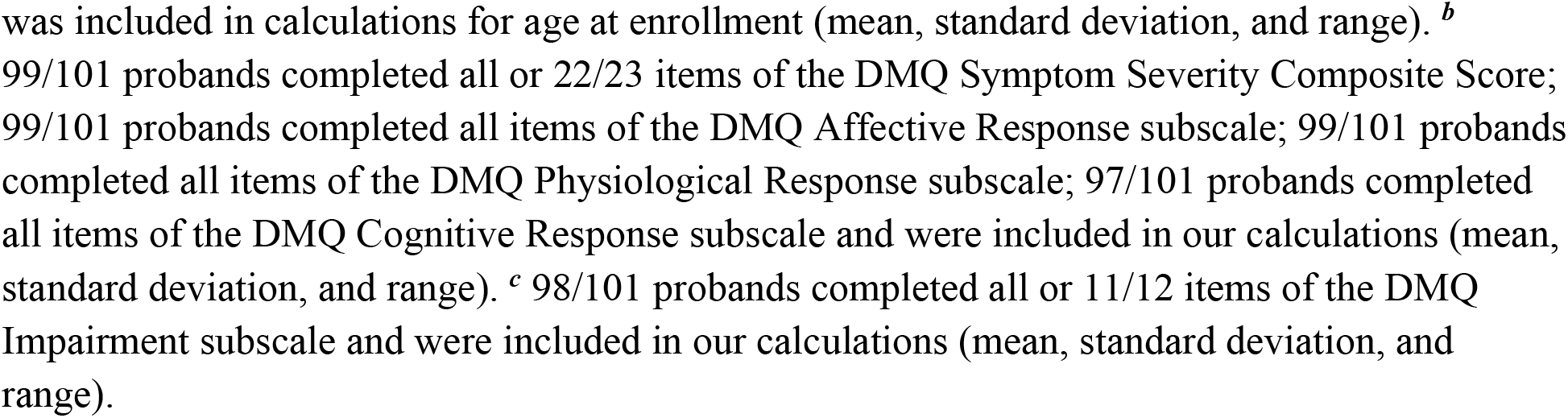
Demographic characteristics and co-occurring conditions among probands with misophonia.

|  | Total (n=101) |
| --- | --- |
| Age, mean (±) s.d. (range) | |
| Age at enrollment | 24.6 ± 11.6 (8-64)<sup>a</sup> |
| Age of onset | 9.52 ± 3.50 (0-22) |
| DMQ scores, mean (±) s.d. (range) | |
| DMQ Symptom Severity Composite Score (> 41 is high) | 63.8 ± 13.6 (28-89)<sup>b</sup> |
| DMQ Affective Response score (max score = 32) | 24.4 ± 5.02 (14-32) |
| DMQ Physiological Response score (max score = 20) | 10.7 ± 4.32 (1-20) |
| DMQ Cognitive Response score (max score = 40) | 28.8 ± 6.43 (15-40) |
| DMQ Impairment score (14-38 is moderate) | 22.1 ± 10.8 (3-46)<sup>c</sup> |
| Sex, n (%) |  |
| Female | 89 (88%) |
| Male | 12 (12%) |
| Race, n (%) |  |
| White | 100 (99%) |
| American Indian/Alaska Native | 1 (1%) |
| Ethnicity, n (%) |  |
| Non-Hispanic/Non-Latino | 93 (92%) |
| Hispanic/Latino | 6 (6%) |
| Unknown | 2 (2%) |
| Self-reported co-occurring conditions, n (%) |  |
| Anxiety | 71 (70%) |
| Depression | 38 (38%) |
| ADHD | 31 (31%) |
| OCD | 25 (25%) |
| Skin picking disorder | 14 (14%) |
| ASD | 11 (11%) |
| Trichotillomania | 6 (6%) |
| Chronic motor/vocal tics | 2 (2%) |
| Tourette disorder | 2 (2%) |
| Intellectual disabilities | 0 |
| Other psychiatric disorders | 6 (6%) |
| Other neurological disorders | 2 (2%) |

Characteristics of 101 probands with self-reported misophonia. DMQ, Duke Misophonia Questionnaire; ADHD, attention-deficit/hyperactivity disorder; OCD, obsessive-compulsive disorder; ASD, autism spectrum disorder. <sup>a</sup> Date of birth was available for 100/101 probands and
was included in calculations for age at enrollment (mean, standard deviation, and range). <sup>b</sup> 99/101 probands completed all or 22/23 items of the DMQ Symptom Severity Composite Score; 99/101 probands completed all items of the DMQ Affective Response subscale; 99/101 probands completed all items of the DMQ Physiological Response subscale; 97/101 probands completed all items of the DMQ Cognitive Response subscale and were included in our calculations (mean, standard deviation, and range). <sup>c</sup> 98/101 probands completed all or 11/12 items of the DMQ Impairment subscale and were included in our calculations (mean, standard deviation, and range).

Overall, individuals with misophonia reported high rates of relatives with misophonia and other co-occurring conditions. Specifically, 39% of probands reported having at least one first-degree relative with misophonia, 65% with anxiety, 57% with depression, and 40% with ADHD (**Table 2**). When considering relatives of any degree, these proportions increased to 48% for misophonia, 71% for anxiety, 63% for depression, and 45% for ADHD (**Table 2**).

**Table 2.**
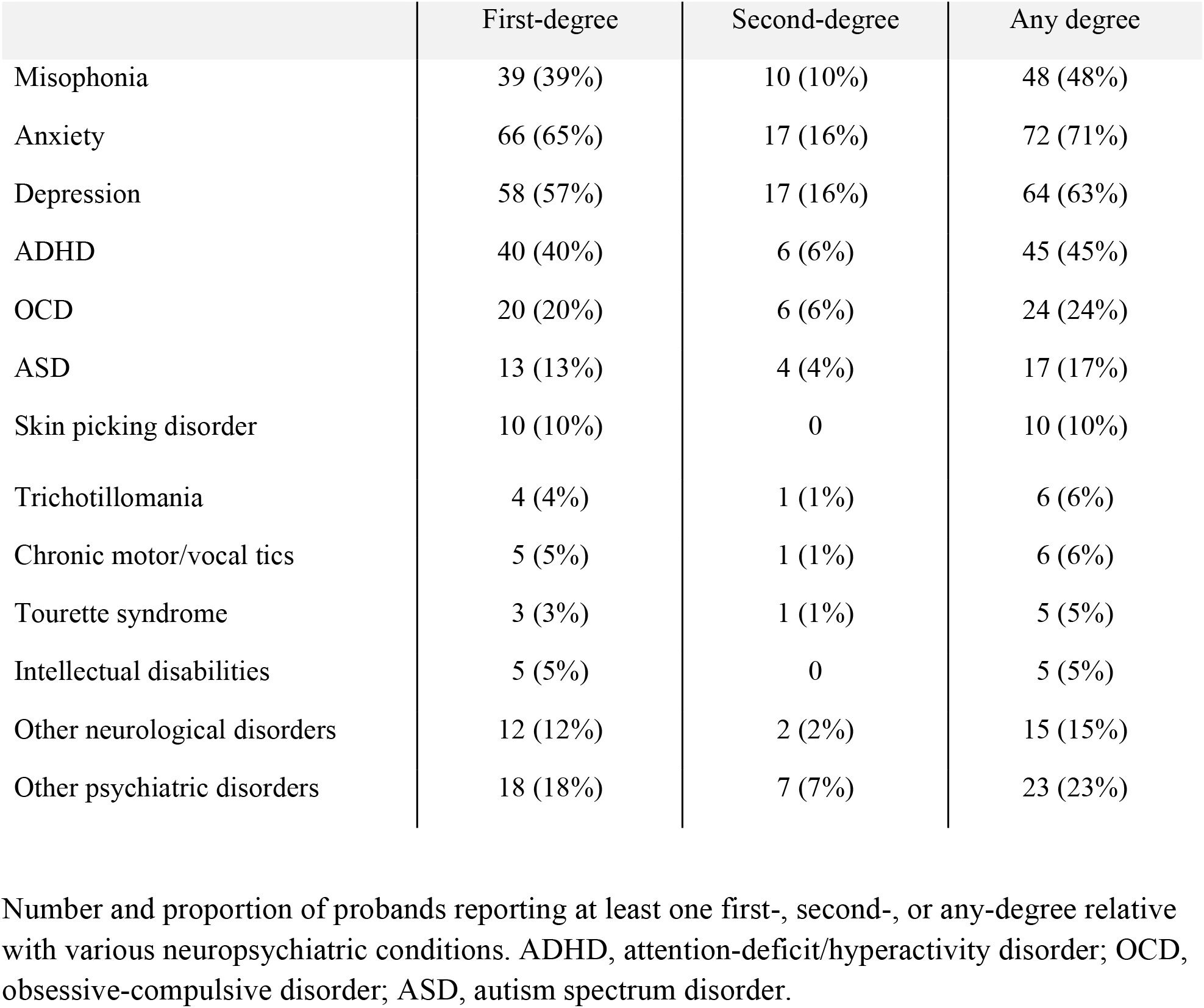
Neuropsychiatric conditions in first-degree, second-degree, or any-degree relatives of probands with misophonia.

Of the 101 probands, 62 were from simplex families and 39 were from multiplex families. The simplex and multiplex probands did not differ significantly in age (at enrollment or onset), sex, race, ethnicity, or co-occurring conditions (**Table S1**).

A comparison of simplex and multiplex family histories revealed no significant differences across most co-occurring conditions, except for misophonia and ADHD among relatives of any degree (**Table S2**). Of the 39 multiplex probands, 31 had at least one parent with misophonia, 13 had a sibling with misophonia, and 1 had a child with misophonia.

Among parents who completed surveys (84 mothers and 81 fathers), 29 parents (24 mothers, five fathers) self-reported a diagnosis of misophonia, and 2 additional fathers reported “unsure” to misophonia and met the threshold values for clinically significant symptoms. Comparing mothers and fathers of probands with misophonia, mothers had significantly higher rates of misophonia (29% vs. 9%; *p* = 0.0012; O.R. = 4.19; 95% C.I. 1.61-12.34) and anxiety (44% vs. 26%; *p* = 0.022; O.R. = 2.24; 95% C.I. 1.11-4.60). No other comparisons were statistically significant (**Figure 1, Table S3**).

**Figure 1:**
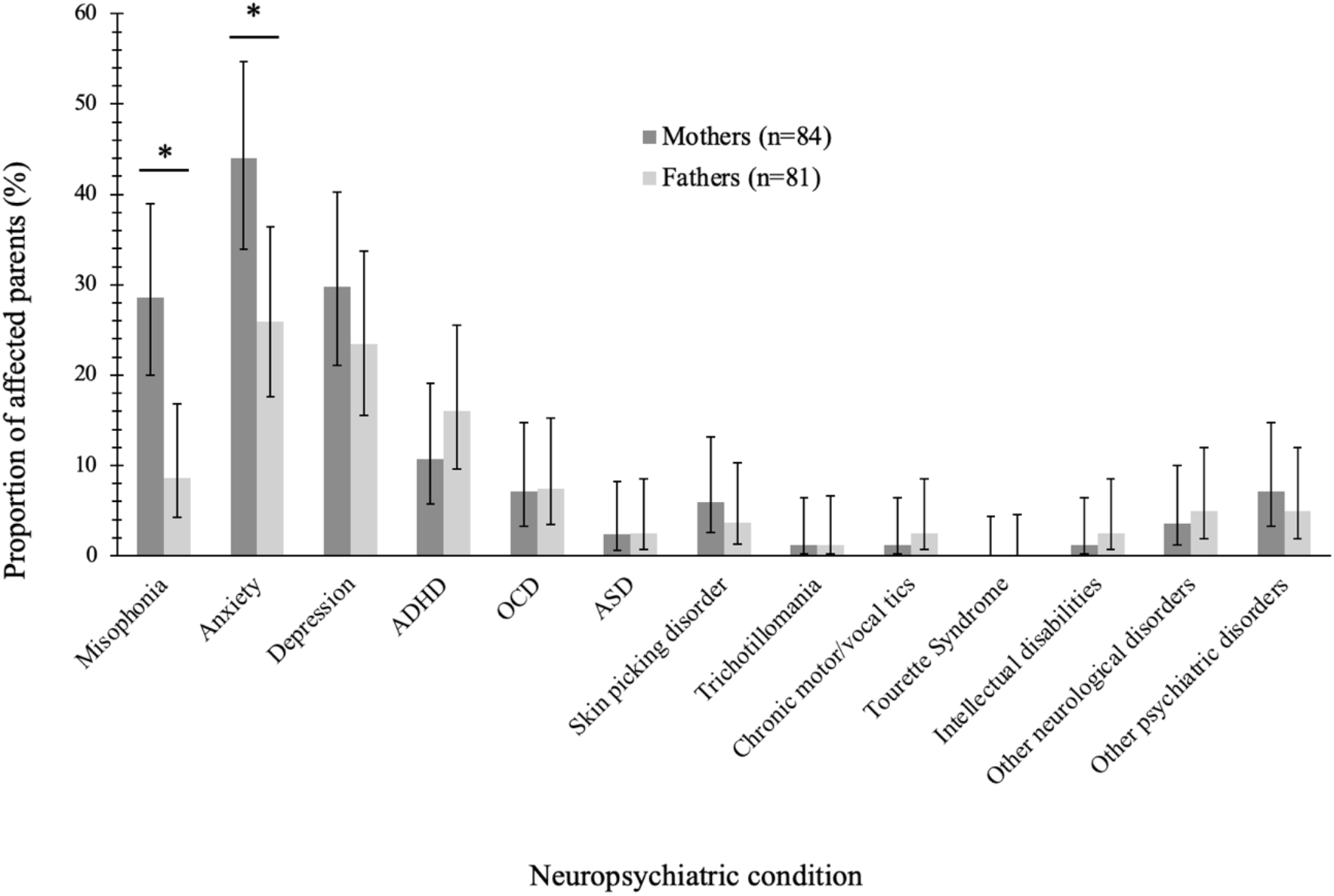
Neuropsychiatric conditions in mothers and fathers of probands with misophonia. A significant difference was found between the rates of mothers versus fathers with self-reported misophonia (n_mothers_ = 24; n_fathers_ = 7; *p* = 0.0012; O.R. = 4.19; 95% C.I. 1.61-12.34) and anxiety (n_mothers_ = 37; n_fathers_ = 21; *p* = 0.022; O.R. = 2.24; 95% C.I. 1.11-4.60) \**p* < 0.05 in two-sided Fisher’s exact test. Error bars represent 95% confidence intervals.

## Discussion

This investigation highlights the high prevalence of misophonia and co-occurring neuropsychiatric conditions among relatives of individuals with misophonia. In this study, 39% of probands with misophonia had at least one first-degree relative with misophonia, and 48% had an affected family member of any degree (**Table 2**). Previous studies of individuals with misophonia have reported a wide range of rates of relatives with similar symptoms (8.33%,^11^ 22%,^1^ 28.2%,^3^ 43.5%,^13^ and 54%^14^). Two studies reported on the degree of relatedness among affected relatives of individuals with misophonia. In a smaller study of 69 individuals with misophonia, 43.5% had a first-degree relative with similar symptoms.^13^ A second study of 90 children and adolescents with misophonia reported that 54% also had a relative with misophonia symptoms, including affected parents (35%), siblings (7%), grandparents (23%), and uncles/aunts (23%).^14^ For context, the estimated prevalence of misophonia in the general population is approximately 5%, though published estimates vary widely by population and assessment method.^16^ The observation that 39% of probands had at least one first-degree relative with misophonia is noteworthy and consistent with familial aggregation, though formal recurrence risk calculations would require a matched control group. Our findings extend prior work by providing estimates of the recurrence rate of misophonia in relatives, underscoring possible shared genetic or environmental predispositions that may contribute to its transmission within families. Unlike prior studies that relied exclusively on proband reports of family history, our study directly surveyed biological parents, providing more reliable estimates of parental misophonia status.

Our results also show that individuals with misophonia have high rates of relatives with other neuropsychiatric conditions. Table 2 highlights that among probands with misophonia, 65% had a first-degree relative with anxiety, 57% with depression, 40% with ADHD, and 13% with autism. For reference, general population prevalence estimates are approximately 18%-31% for anxiety disorders,^17,18^ 8-13% for depression,^19,20^ and 4-6% for ADHD^21,22^ in U.S. adults. However, differences in ascertainment methods, demographics, and diagnostic definitions between our sample and these population surveys preclude direct comparison, and a matched control group would be needed to determine whether the rates observed here are significantly elevated. The rates of neuropsychiatric conditions among relatives also mirror the high rates of these conditions observed in the probands (Table 1) and align with previous research demonstrating a significant overlap between misophonia and several neuropsychiatric conditions.^16,23-27^ Past research supports significant genetic correlations between misophonia and major depressive disorder, post-traumatic stress disorder, and generalized anxiety disorder^28^ as well as strong associations to neuroticism.^28,29^ A study of mothers of youth with misophonia showed significantly higher rates of postpartum depression in mothers of affected children than controls,^14^ suggesting a potential shared predisposition. Neurobiological studies of misophonia suggest disruptions in salience detection and emotional processing networks,^30^ heightened sensory sensitivity,^31^ and emotion dysregulation,^32^ which are established transdiagnostic risk factors across anxiety, mood, and neurodevelopmental disorders, suggesting potential shared biological mechanisms. Importantly, some of these co-occurring conditions, particularly anxiety and depression, may also develop as consequences of living with misophonia (e.g., anxiety about encountering triggers, or depression related to social isolation from avoidance behaviors), rather than representing independent co-occurring conditions. Alternatively, elevated family history rates of neuropsychiatric conditions may serve as a stressor contributing to misophonia development.^14,32^ Additional research is needed to investigate the directionality of these relationships, and longitudinal studies may shed light on early development and familial predispositions to misophonia.

A more detailed investigation of parental family history of misophonia suggests that affected probands have a higher likelihood of having an affected mother than an affected father. Mothers had significantly higher rates of misophonia (29% vs. 9%) and anxiety (44% vs. 26%) than fathers (**Figure 1**). These higher rates of affected mothers than fathers mirror previous work by Rouw and Erfanian^1^ who found that individuals with misophonia reported having more mothers (n=48/301) than fathers (n=19/301) with similar misophonia symptoms. While this previous study relied on proband self-reports of their parents’ misophonia symptoms, our study used parental reports of their own histories, which may reduce reporting bias. The observed excess of affected mothers over fathers is consistent with different genetic models, though the known female preponderance of both misophonia and anxiety could also account for this pattern.

Our results are also consistent with a female preponderance of misophonia. In this study, 88% of affected probands were female (**Table 1**), and more mothers (29%) than fathers (9%) were affected with misophonia (**Figure 1**). The female preponderance of misophonia is well supported in past research,^1,3,5,10,13^ with a minority of studies reporting no significant sex difference.^11,12^ The mechanisms underlying this difference remain unclear. Possible explanations include sex differences in sensory processing or emotional reactivity, hormonal influences, differential symptom expression or recognition, or ascertainment biases if females are more likely to seek support or participate in research. Notably, the high rate of co-occurring anxiety in our sample is consistent with anxiety disorders being more prevalent in females,^33,34^ raising the question of whether the female predominance in misophonia reflects, in part, shared vulnerability pathways with anxiety disorders.

In exploratory analyses, we compared clinical characteristics of probands from simplex and multiplex families to assess possible differences in presentation between those with a family history of misophonia and those without. Among these probands, no significant differences were found in clinical characteristics, including misophonia severity and impairment, or in the presence of co-occurring conditions (**Table S1**). When assessing the family history of other neuropsychiatric conditions, we generally found no significant differences between simplex and multiplex families, though multiplex probands had significantly higher rates of any-degree relatives with ADHD than simplex probands (59% vs. 35%, *p*<0.05) (**Table S2**).

Several limitations of this study should be considered. First, our study may be subject to self-selection bias, as participants with a family history of misophonia may have been more inclined to enroll, possibly due to greater awareness of the familial nature of their condition. This bias is expected to inflate our estimates of familial recurrence. Of note, the 39% first-degree familial rate observed here falls within the wide range previously reported (8-54%), and is lower than the 54% reported by Siepsiak et al. in a clinic-based pediatric sample,^14^ suggesting that our estimates are not inconsistent with those obtained through other ascertainment methods. Nonetheless, readers should interpret our familial recurrence rates as upper-bound estimates that may overestimate the true rate in the broader misophonia population. Second, our data rely on self-reported diagnoses, family history, and measures, which may be subject to recall bias or underreporting due to limited awareness, insight, or stigma. Direct interviews and structured diagnostic assessments of misophonia and all co-occurring conditions may yield more precise and potentially lower prevalence estimates. Third, our study had limited racial, ethnic, and sex diversity. As previously mentioned, most of our participants self-identified as white (99%) and female (88%). Future efforts should be made to increase enrollment of underrepresented groups.

Fourth, our sample size was modest (n=101). Future studies should include a larger, more diverse sample to ensure that findings are generalizable. Fifth, our study lacked a control group of individuals without misophonia, limiting our ability to determine whether the observed rates of co-occurring conditions and family history are truly elevated compared to controls. As such, our findings describe patterns of familial aggregation rather than formally demonstrating familiality in the genetic epidemiological sense. Sixth, recruitment through online misophonia support groups likely enriched our sample for individuals with more severe symptoms. Indeed, the mean DMQ Severity Score in our sample (63.8) substantially exceeded the threshold for high symptoms (>41), suggesting our findings may not generalize to individuals with milder presentations. Seventh, given the exploratory nature of this study and the number of statistical comparisons performed, we did not correct for multiple comparisons. Some nominally significant findings may represent false positives, and results should be interpreted cautiously pending replication. Finally, the cross-sectional design precludes causal inference about the relationship between family history and the development of misophonia. Despite these limitations, our study provides new and important insights into the clinical characteristics and family history patterns of misophonia.

## Conclusion

In conclusion, this study provides insight into the familial patterns of misophonia. We demonstrate the high rates of misophonia and other neuropsychiatric conditions in family members, suggesting possible shared genetic and environmental risk factors. We also found higher rates of misophonia and anxiety among mothers than fathers, consistent with the female predominance of these conditions. Interestingly, the clinical features, severity, and functional impairment between affected probands were similar regardless of simplex or multiplex status. While these findings require replication in larger, more diverse samples with control groups, they provide a foundation for future genetic and environmental studies of misophonia etiology and suggest that clinicians evaluating patients with misophonia may benefit from inquiring about family history of sound sensitivity and neuropsychiatric conditions. Together, these results provide new insight into the features and familial patterns of misophonia, generating hypotheses about possible heritable factors that may inform future intervention strategies and enhance understanding of the disorder.

## Data Availability

All data produced in the present study are available upon reasonable request to the authors.

## Acknowledgements

This paper was reviewed by the soQuiet Lived Experience Advisory Panel [LEAP] for inclusion of accurate and earnest representations of people who personally experience misophonia and related conditions.

