## Supplemental Information for "Family history of misophonia and co-occurring neuropsychiatric conditions"

**Supplementary Information for "Family history of misophonia and co-occurring neuropsychiatric conditions"**

File Includes:

Supplementary Tables 1-3

**Table S1:** Demographic characteristics and co-occurring conditions in simplex and multiplex probands with misophonia

| | **Total (n=101)** | **Simplex (n=62)** | **Multiplex (n=39)** | ***p*-value** |
|---|---|---|---|---|
| Age, mean (±) s.d. (range) | | | | |
| Age at enrollment | 24.6 ± 11.6 (8-64)[a] | 24.9 ± 11.7 (8-51) | 23.7 ± 12.0 (8-64) | 0.63 |
| Age of onset | 9.52 ± 3.50 (0-22) | 9.30 ± 3.42 (0-19) | 9.88 ± 3.69 (5-22) | 0.45 |
| DMQ scores, mean (±) s.d. (range) | | | | |
| DMQ Symptom Severity Composite Score (> 41 is high) | 63.8 ± 13.6 (28-89)[b] | 64.2 ± 14.2 (28-88)[d] | 63.3 ± 12.8 (36-89)[f] | 0.75 |
| DMQ Affective Response score (max score = 32) | 24.4± 5.02 (14-32) | 24.4 ± 5.00 (14-32) | 24.4 ± 5.12 (14-32) | 0.99 |
| DMQ Physiological Response score (max score = 20) | 10.7 ± 4.32 (1-20) | 10.8 ± 4.37 (1-20) | 10.6 ± 4.29 (4-18) | 0.85 |
| DMQ Cognitive Response score (max score = 40) | 28.8 ± 6.43 (15-40) | 29.1 ± 6.79 (15-40) | 28.3 ± 5.84 (17-40) | 0.53 |
| DMQ Impairment score (14-38 is moderate) | 22.1 ± 10.8 (3-46)[c] | 22.5 ± 11.4 (3-46)[e] | 21.4 ± 9.81 (4-43)[g] | 0.63 |
| Gender, n (%) | | | | |
| Female | 89 (88%) | 55 (89%) | 34 (87%) | 1 |
| Male | 12 (12%) | 7 (11%) | 5 (13%) | 1 |
| Race, n (%) | | | | |
| White/European Caucasian | 100 (99%) | 61 (98%) | 39 (100%) | 1 |
| American Indian/Alaska Native | 1 (1%) | 1 (2%) | 0 | 1 |
| Ethnicity, n (%) | | | | |
| Non-Hispanic/Non-Latino | 93 (92%) | 57 (92%) | 36 (92%) | 1 |
| Hispanic/Latino | 6 (6%) | 4 (6%) | 2 (5%) | 1 |
| Self-reported comorbid disorders, n (%) | | | | |
| Anxiety | 71 (70%) | 47 (76%) | 24 (62%) | 0.18 |
| Depression | 38 (38%) | 26 (42%) | 12 (31%) | 0.30 |
| ADHD | 31 (31%) | 19 (31%) | 12 (31%) | 1 |
| OCD | 25 (25%) | 15 (24%) | 10 (26%) | 1 |
| Skin picking disorder | 14 (14%) | 10 (16%) | 4 (10%) | 0.56 |
| ASD | 11 (11%) | 6 (10%) | 5 (13%) | 0.75 |
| Trichotillomania | 6 (6%) | 3 (5%) | 3 (8%) | 0.67 |
| Other psychiatric disorders | 6 (6%) | 2 (3%) | 4 (10%) | 0.20 |
| Chronic motor/vocal tics | 2 (2%) | 0 | 2 (5%) | 0.15 |
| Tourette Syndrome | 2 (2%) | 2 (3%) | 0 | 0.52 |

| | | | | |
|---|---|---|---|---|
| Other neurological disorders | 2 (2%) | 1 (2%) | 1 (3%) | 1 |
| Intellectual disabilities | 0 | 0 | 0 | 1 |

Characteristics of 62 simplex probands and 39 multiplex probands with self-reported misophonia. Probands were considered from multiplex families if they had at least one first-degree relative with misophonia and simplex if no other first-degree relatives were affected. No significant differences were found for any characteristics between groups. DMQ, Duke Misophonia Questionnaire; ADHD, attention-deficit/hyperactivity disorder; OCD, obsessive-compulsive disorder; ASD, autism spectrum disorder. [a] Date of birth available for 100/101 probands and was included in calculations for age at enrollment (mean, standard deviation, and range). [b] 99/101 probands completed all or 22/23 items of the DMQ Symptom Severity Composite Score; 99/101 probands completed all items of the DMQ Affective Response subscale; 99/101 probands completed all items of the DMQ Physiological Response subscale; 97/101 probands completed all items of the DMQ Cognitive Response subscale and were included in our calculations (mean, standard deviation, and range). [c] 98/101 probands completed all or 11/12 items of the DMQ Impairment subscale and were included in our calculations (mean, standard deviation, and range). [d] 61/62 probands completed all or 22/23 items of the DMQ Symptom Severity Composite Score; 60/62 probands completed all items of the DMQ Affective Response subscale; 61/62 probands completed all items of the DMQ Physiological Response subscale; 60/62 probands completed all items of the DMQ Cognitive Response subscale and were included in our calculations (mean, standard deviation, and range). [f] 38/39 probands completed all or 22/23 items of the DMQ Symptom Severity Composite Score; 38/39 probands completed all items of the DMQ Physiological Response subscale; 37/39 probands completed all items of the DMQ Cognitive Response subscale and were included in our calculations (mean, standard deviation, and range). [g] 37/39 probands completed all or 11/12 items of the DMQ Impairment subscale and were included in our calculations (mean, standard deviation, and range).

**Table S2:** Neuropsychiatric conditions in first-degree, second-degree, or any-degree relatives in simplex and multiplex probands with misophonia.

| | First-degree | | | | Second-degree | | | | Any degree | | | |
|---|---|---|---|---|---|---|---|---|---|---|---|---|
| | **Multiplex (n=39)** | **Simplex (n=62)** | ***p*-value** | **Odds ratio** | **Multiplex (n=39)** | **Simplex (n=62)** | ***p*-value** | **Odds ratio** | **Multiplex (n=39)** | **Simplex (n=62)** | ***p*-value** | **Odds ratio** |
| Misophonia | 39 | 0 | **-** | **-** | 3 (8%) | 7 (11%) | 0.74 | 0.657 | 39 | 9 (15%) | **<.001** | Inf |
| Anxiety | 29 (74%) | 38 (61%) | 0.20 | 1.82 | 5 (13%) | 12 (19%) | 0.43 | 0.616 | 30 (77%) | 43 (69%) | 0.50 | 1.47 |
| Depression | 27 (69%) | 31 (50%) | 0.066 | 2.23 | 5 (13%) | 12 (19%) | 0.43 | 0.616 | 27 (69%) | 37 (59%) | 0.40 | 1.42 |
| ADHD | 20 (51%) | 20 (32%) | 0.064 | 2.19 | 3 (8%) | 3 (5%) | 0.67 | 1.63 | 23 (59%) | 22 (35%) | **<0.05** | 2.59 |
| OCD | 9 (23%) | 11 (18%) | 0.61 | 1.39 | 1 (3%) | 5 (8%) | 0.40 | 0.303 | 9 (23%) | 15 (24%) | 1 | 0.941 |
| Skin picking disorder | 6 (15%) | 4 (6%) | 0.18 | 2.61 | 0 | 0 | 1 | 0 | 6 (15%) | 4 (6%) | 0.30 | 2.61 |
| ASD | 6 (15%) | 7 (11%) | 0.56 | 1.42 | 0 | 4 (6%) | 0.16 | 0 | 7 (18%) | 10 (16%) | 0.79 | 1.14 |
| Trichotillomania | 3 (8%) | 1 (2%) | 0.30 | 5.00 | 0 | 1 (2%) | 1 | 0 | 3 (8%) | 3 (5%) | 0.67 | 1.63 |
| Chronic motor/vocal tics | 2 (5%) | 3 (5%) | 1 | 1.06 | 1 (3%) | 0 | 0.39 | Inf | 3 (8%) | 3 (5%) | 0.67 | 1.63 |
| Tourette syndrome | 1 (3%) | 2 (3%) | 1 | 0.791 | 0 | 1 (2%) | 1 | 0 | 2 (5%) | 3 (5%) | 1 | 1.06 |
| Intellectual disabilities | 4 (10%) | 1 (2%) | 0.072 | 6.84 | 0 | 0 | 1 | 0 | 4 (10%) | 1 (2%) | 0.072 | 6.84 |
| Other neurological disorders | 5 (13%) | 7 (11%) | 1 | 1.15 | 0 | 2 (3%) | 0.52 | 0 | 6 (15%) | 9 (15%) | 1 | 1.07 |
| Other psychiatric disorders | 7 (18%) | 11 (18%) | 1 | 1.01 | 1 (3%) | 6 (10%) | 0.24 | 0.248 | 7 (18%) | 16 (26%) | 0.47 | 0.632 |

Number and proportion of multiplex (n=39) and simplex (n=62) probands who reported having at least one first-, second-, or any-degree relative with self-reported neuropsychiatric conditions. A significant difference was found between multiplex and simplex in the proportion of any-degree relatives with misophonia ($p$ = 1.14e-19; O.R. = Inf) and ADHD ($p$ = 0.025; O.R. = 2.59). p-values were calculated using a two-sided Fisher's exact test. Bold represents p-values < 0.05.

**Table S3:** Neuropsychiatric conditions in mothers and fathers of probands with misophonia.

| | Mothers (n=84) | Fathers (n=81) | *p*-value | Odds ratio |
|---|---|---|---|---|
| Misophonia | 24 (28.6%) | 7 (8.64%) | **0.0013** | 4.19 |
| Anxiety | 37 (44.0%) | 21 (25.9%) | **0.022** | 2.24 |
| Depression | 25 (29.8%) | 19 (23.5%) | 0.38 | 1.38 |
| ADHD | 9 (10.7%) | 13 (16.0%) | 0.36 | 0.629 |
| OCD | 6 (7.14%) | 6 (7.41%) | 1 | 0.962 |
| Skin picking disorder | 5 (5.95%) | 3 (2.70%) | 0.72 | 1.64 |
| ASD | 2 (2.38%) | 2 (2.47%) | 1 | 0.964 |
| Trichotillomania | 1 (1.19%) | 1 (1.23%) | 1 | 0.964 |
| Chronic motor/vocal tics | 1 (1.19%) | 2 (2.47%) | 0.62 | 0.478 |
| Tourette syndrome | 0 | 0 | 1 | 0 |
| Other psychiatric disorders | 6 (7.14%) | 4 (4.94%) | 0.75 | 1.48 |
| Other neurological disorders | 3 (3.57%) | 4 (4.94%) | 0.72 | 0.714 |
| Intellectual disabilities | 1 (1.19%) | 2 (2.47%) | 0.62 | 0.478 |

Number and proportion of self-reported neuropsychiatric conditions in parents of probands with misophonia. A significant difference was found in the rates of mothers versus fathers who self-reported being affected by misophonia ($n_{mothers} = 24$; $n_{fathers} = 7$; $p = 0.0013$; O.R. = 4.19; 95% C.I. 1.61-12.34) and anxiety ($n_{mothers} = 37$; $n_{fathers} = 21$; $p = 0.022$; O.R. = 2.24; 95% C.I. 1.11-4.60). Bold values represent $p < 0.05$.
